# Distributed Genetic Effects of the Corpus Callosum Subregions Suggest Links to Neuropsychiatric Disorders and Related Traits

**DOI:** 10.1101/2023.05.13.23289938

**Authors:** Megan L. Campbell, Shareefa Dalvie, Alexey Shadrin, Dennis van der Meer, Kevin O’Connell, Olekasander Frei, Ole A. Andreassen, Dan J. Stein, Jaroslav Rokicki

## Abstract

**Background:** The corpus callosum (CC) is a brain structure with a high heritability and potential role in psychiatric disorders. However, the genetic architecture of the CC and the genetic link with psychiatric disorders remains largely unclear. We investigated the genetic architectures of the volume of the CC and its subregions, and the genetic overlap with psychiatric disorders.

**Methods:** We applied multivariate GWAS to genetic and T1-weighted MRI data of 40,894 individuals from the UK Biobank, aiming to boost genetic discovery and to assess the pleiotropic effects across volumes of the five subregions of the CC (posterior, mid posterior, central, mid anterior and anterior) obtained by FreeSurfer 7.1. Multivariate GWAS was run combining all subregions, co-varying for relevant variables. Gene-set enrichment analyses were performed using MAGMA. Linkage disequilibrium score regression (LDSC) was used to determine SNP-based heritability of total CC volume and volumes of its subregions as well as their genetic correlations with relevant psychiatric traits.

**Results:** We identified 70 independent loci with distributed effects across the five subregions of the CC (p < 5 × 10^−8^). Additionally, we identified 33 significant loci in the anterior subregion, 23 in the mid anterior, 29 in the central, 7 in the mid posterior and 56 in the posterior subregion. Gene-set analysis revealed 156 significant genes contributing to volume of the CC subregions (p < 2.6 × 10^−6^). LDSC estimated the heritability of CC to (h^2^_SNP_=0.38, SE=0.03), and subregions ranging from 0.22 (SE=0.02) to 0.37 (SE=0.03). We found significant genetic correlations of total CC volume with bipolar disorder (BD, rg=-0.09, SE=0.03; p=5.9 × 10^−3^) and drinks consumed per week (rg=-0.09, SE=0.02; p=4.8 × 10^−4^), and volume of the mid anterior subregion with BD (rg=-0.12, SE=0.02; p=2.5 × 10^−4^), major depressive disorder (rg=-0.12, SE=0.04; p=3.6 × 10^−3^), drinks consumed per week (rg=-0.13, SE=0.04; p=1.8 × 10^−3^) and cannabis use (rg=-0.09, SE=0.03; p=8.4 × 10^−3^).

**Conclusions:** Our results demonstrate that the CC has a polygenic architecture implicating multiple genes, and show that CC subregion volumes are heritable. We found distinct genetic factors are involved in the development of anterior and posterior subregions, consistent with their divergent functional specialization. Significant genetic correlation between volumes of the CC and bipolar disorder, drinks per week, major depressive disorder and cannabis consumption subregion volumes with psychiatric traits is noteworthy and deserving of further investigation.

## Introduction

The corpus callosum (CC) is the largest white matter tract in the brain, and serves as the major interhemispheric commissure, integrating information from the two cerebral hemispheres to facilitate language, affective and sensorimotor function (Wang et al., 2020). The CC is commonly segmented into five subregions: anterior, mid-anterior, central, mid-posterior and posterior, with each subregion having distinct axonal projections from cortical regions (Hofer and Frahm, 2006). For example, projections from the auditory and somatosensory cortices primarily cross the central subregion, while those from the visual cortex cross the anterior subregion (Hofer and Frahm, 2006). Although the exact boundaries of these subregions are challenging to delineate using MRI (Hofer and Frahm, 2006), their functional importance in various cognitive processes such as language, sensory processing, and emotional regulation is well-established (Cover *et al*., 2018).

Differences in volume of the CC have been associated with several psychiatric disorders including mood, substance use and developmental disorders (Rosenbloom, Sullivan and Pfefferbaum, 2003; Newbury and Rosen, 2012; Abramovic et al., 2018; Prunas et al., 2018; de Souza et al., 2019). Volumetric reductions in the CC have been observed in bipolar disorder (BD) patients, with their relatives showing intermediate volumetric reductions between controls and BD probands (Walterfang et al., 2009; Francis et al., 2016). There is evidence to suggest that alcohol consumption affects CC morphometry (de Souza et al., 2019). These effects appear to be subregion-specific, with volume of the anterior CC subregion displaying a negative association with alcohol consumption, irrespective of an alcohol use disorder diagnosis (de Souza et al., 2019). Frontal and superior subregions of the CC appear to be most affected by alcohol consumption, with the inferior and posterior CC subregions remain relatively preserved (de Souza et al., 2019). CC volume reductions are also present in autism spectrum disorder (ASD), with the anterior and mid anterior subregions seemingly most affected (Frazier and Hardan, 2009; Valenti *et al*., 2020). Differences in volume of the CC subregions have also been noted in attention deficit hyperactivity disorder (ADHD), major depressive disorder (MDD) and schizophrenia (SCZ), although the pattern of alteration in these disorders is less clear (Connaughton *et al*., 2022; Zhao *et al*., 2022; Zhou *et al*., 2022).

Twin studies have demonstrated a high heritability of CC size, estimated to 0.67 for total CC and 0.48-0.62 for the subregions (Scamvougeras et al., 2003; Woldehawariat et al., 2014). Although twin and family studies have established the heritability of the CC, there has been a lack of large-scale genetic investigations aimed at identifying the contribution of common genetic variants to the morphology of CC. Recent advances in methods for exploring the genetic architecture of the brain have provided insight into the subregion-specific genetic aetiology of multiple brain regions (van der Meer, Rokicki, *et al*., 2020; Zhao *et al*., 2020; Mufford *et al*., 2021; Bahrami *et al*., 2022; Ou *et al*., 2023). This work has been valuable in identifying shared and specific genetic loci and pathways across subregions, as well as providing insight into differential genetic overlap with psychiatric disorders. Accordingly, we aimed to determine the genetic influences on CC subregions and total CC volume, assess the heritability of this structure, and investigate the genetic correlation with psychiatric traits and disorders.

## Methods

### Processing of Imaging and Genetic Data

We accessed raw, T1-weighted MRI data from 35,592 genotyped, Caucasian individuals of British ancestry (age range: 45 to 81 years; mean age: 64.3 years, SD: 7.5; 52.3% female) from the UK Biobank (Bycroft et al., 2018) (accession code 27412) for our main analysis, and 5,302 individuals of non-Caucasian ancestry for our generalization analyses conducted in independent data (age range: 45 to 81 years; mean age: 62.9, SD: 7.7 years; 46.3% female). Two different methods were used to segment the CC: FreeSurfer v7.1, which is based on the Witelson segmentation (Witelson, 1989), and the division by Hofer and Frahm (2006) implemented with C8 software (Herron, Kang, and Woods, 2012). Both methods divide the CC into five subregions, the differences are illustrated and described in more detail in the Supplementary Material (Supplementary Material, Figure 1). However, the main difference being that FreeSurfer extracts volume of the CC and each subregion while the C8 software extracts thickness measurements along the medial line in the sagittal plane. Given the widespread use of FreeSurfer for subcortical segmentation, we chose to focus on this method and use the approach proposed by Hofer and Frahm (2006) to validate our findings. To account for possible confounders, neuroCombat was used to pre-residualize for age, age squared, sex, scanning site, Euler score (a proxy of image quality), total intracranial volume (ICV) and the first 20 genetic principal components for each CC subregion and total CC volume (Fortin et al., 2018). These variables were included as covariates in the univariate GWAS. The pre-residualization step was performed for both segmentation methods and for both the European and non-European participants. These residuals were used as the outcome variables in subsequent multivariate analyses.

**Figure 1.**
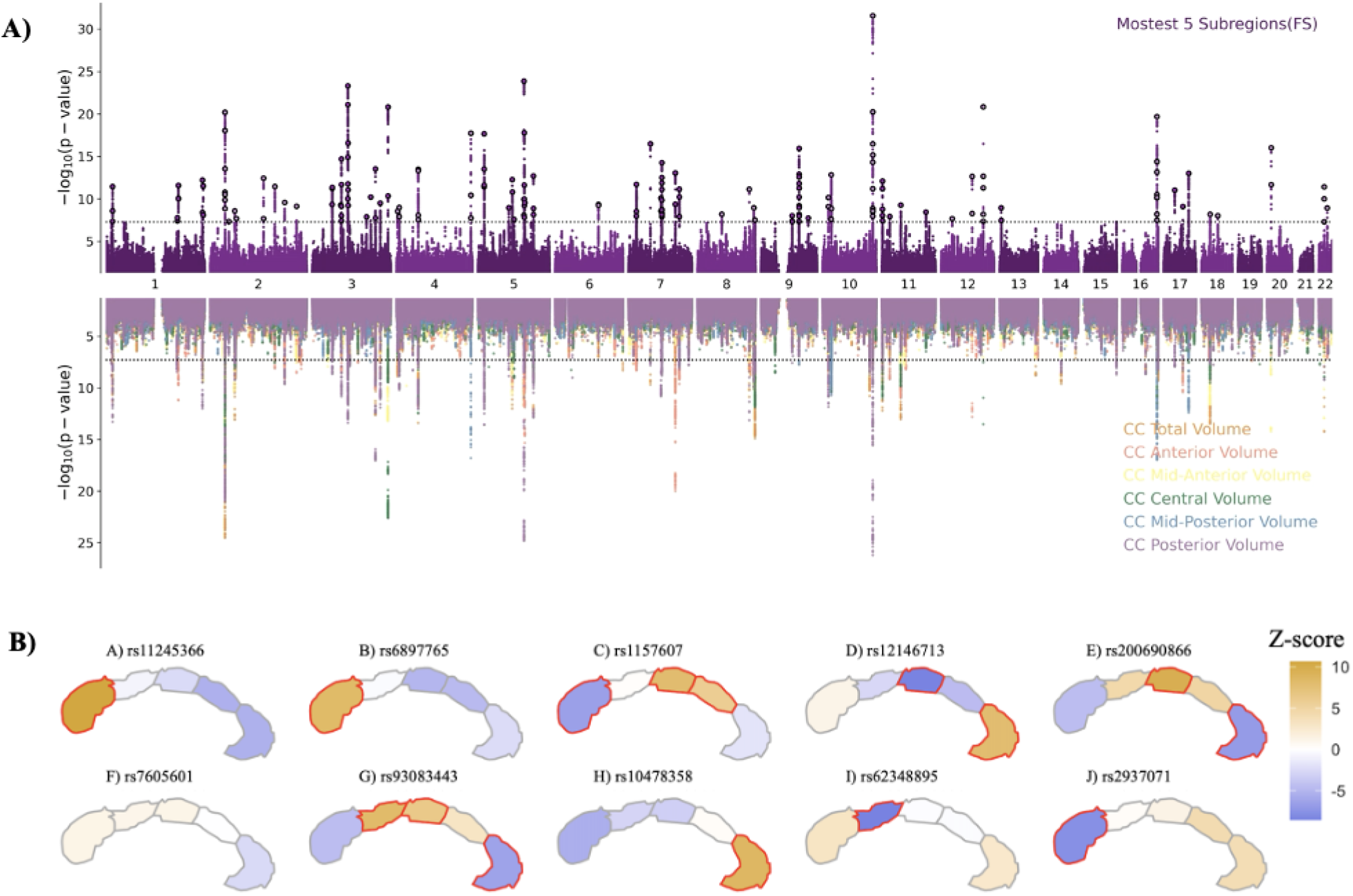
Miami plot of the −log10(p) statistics from the multivariate (top) and univariate (bottom) GWAS of total CC volume. **A)** Top: The multivariate analysis was conducted using MOSTest and illustrate the distributive genetic architecture of the CC. Bottom: Individual univariate GWAS were conducted for total CC volume and each subregion, indicating both a shared and unique genetic etiology of the CC subregions. **B) Top SNPs from the multivariate GWAS show subregion specific effects across the CC**. The top ten SNPs from the multivariate GWAS were mapped to the CC subregions using the Z-scores from the respective univariate subregion GWAS. The results indicate a gradient of effect across the posterior and anterior subregions.

Standard quality control (QC) procedures were used to process the UK Biobank version 3, imputed genetic data: SNPs with an imputation score of less than 0.5, a minor allele frequency (MAF) of less than 0.005, a missingness rate of more than 10% and those that failed the Hardy-Weinberg test of equilibrium (HWE, p<1×10^−9^) were removed. A total of 35,593 individuals remained in the European cohort after QC and an additional 5,286 in the non-European cohort.

### Genome-wide association analyses: Multivariate and Univariate

The pre-residualized CC subregions were used as the outcome in the multivariate analysis. This was conducted using the multivariate omnibus statistical test (MOSTest), which has been previously applied to a variety of brain morphology traits (van der Meer, Frei, *et al*., 2020; Bahrami *et al*., 2022). MOSTest implements permutation testing to identify genetic effects across multiple phenotypes, resulting in a multivariate GWAS summary statistic across all five CC subregions. Replication of the MOSTest results was conducted using MOSTEST-PolyVertex Score (MOSTEST-PVS) (Loughnan et al., 2022) in the non-European sample of 5,210 individuals.

To identify the distinct genetic contribution of every CC subregion, an individual univariate GWAS of each subregion and total CC were performed using Plink v2 (Chang et al., 2015). This approach allows for the determination of the effect direction and enables further follow-up analyses. Additionally, univariate GWAS of the subregions from the Hofer and Frahm segmentation were conducted as an additional replication step. Significant SNPs were defined using a significance threshold of *p*<9.6×10^−9^ (Bonferroni, accounting for 5 subregions and total CC). Independent significant genetic variants are identified as variants with p < 8.3×10^−9^ and LD r^2^<0.6 with each other. For each independent significant variant all candidate variants are identified as variants with LD r2≥0.6 with the lead variant. For a given independent significant variant the borders of the genomic locus are defined as min/max positional coordinates over all corresponding candidate variants. Loci are then merged if they are separated by less than 250kb. A subset of independent significant variants with LD r2<0.1 are then selected as lead variants. If a locus contains several lead SNPs using this definition, then the one with the most significant p-value is selected as a lead SNP representing the locus. In order to identify the unique genetic contribution to each CC subregion, additional univariate GWAS of each subregion were conducted, including total CC volume as a covariate. An overview of the GWAS are provided in the supplementary material, Figure 2.

**Figure 2:**
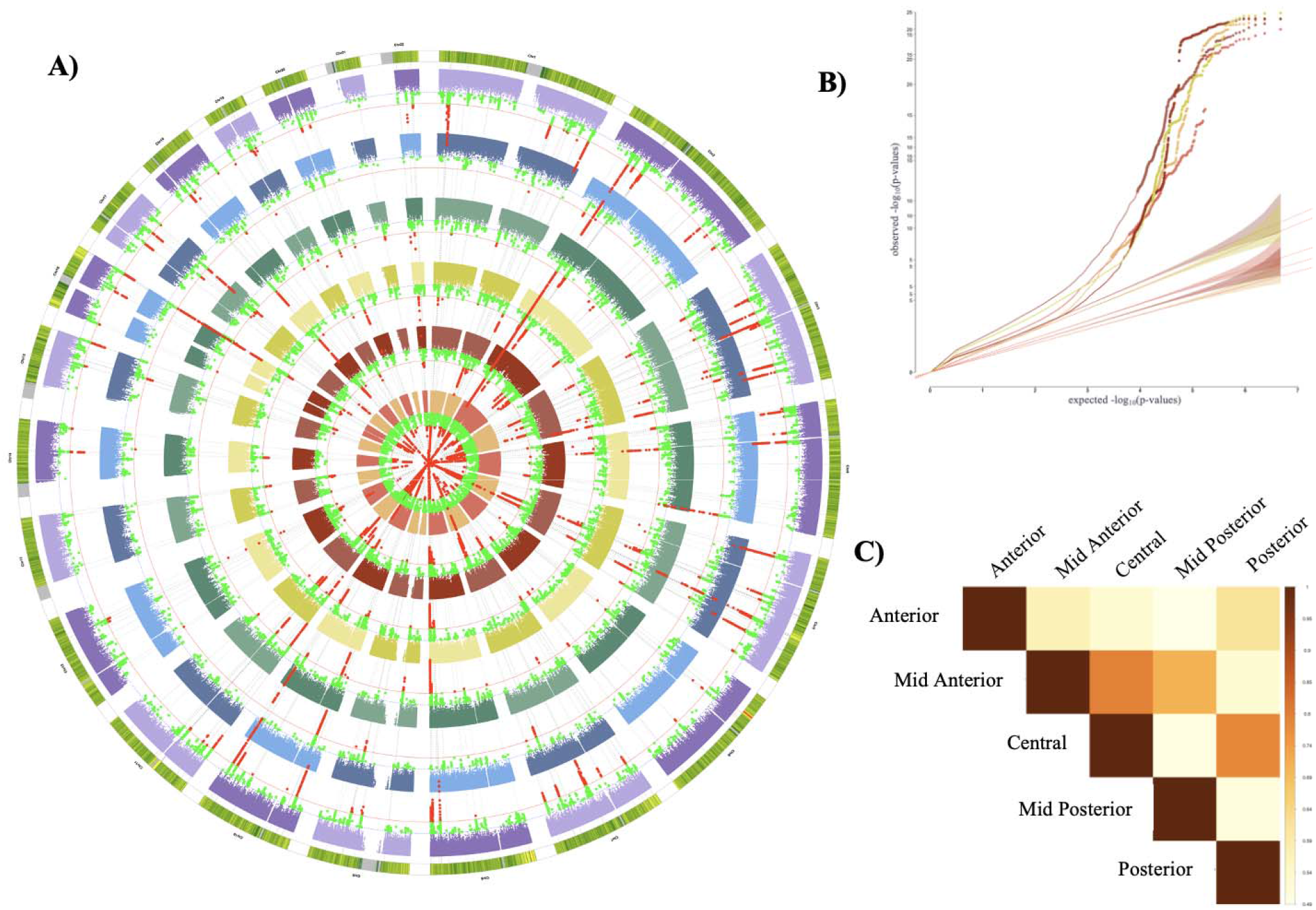
A) Manhattan plots from univariate GWAS of each CC subregion B) Quantile-Quantile plot (QQ plot) of observed versus expected −log_10_(p-values) the GWAS of CC subregions. The shaded regions represent the expected null distribution of −log_10_(p-values) under the assumption of no association. C) Volumetric correlations of CC subregions

### Functional annotation

Functional Mapping and Annotation of Genome-wide Association Studies (FUMA, platform v1.3.7, accessed September 2022) was used to annotate the GWAS results, using the 1000GPhase3 EUR as reference panel and default parameters (Watanabe et al., 2017).We conducted genome-wide gene-based association and gene-set analyses using MAGMA v.1.08 (http://ctg.cncr.nl/software/magma) in FUMA. All variants in the major histocompatibility complex (MHC) region (GRCh37: 6:26,000,000–33,000,000) were excluded before running the MAGMA analyses. MAGMA performs multiple linear regression to map the input SNPs to 18,091 protein coding genes and estimates the significance value of each gene. Genes were considered significant if *p*<2.76×10^−6^ (Bonferroni correction). These genes were investigated for enrichment of biological processes, tissue, and cell types. In addition to FUMA, we used Open Target Genetics which integrates functional and biological data from multiple sources to highlight functionally implicated genes, conduct phenome-wide association studies and tests for association with previous GWAS (Mountjoy *et al*., 2021).

### SNP-Based Heritability and Genetic Correlation

SNP-based heritability (h^2^_SNP_) and genetic correlation were calculated using Linkage Disequilibrium Score Regression (LDSC) (B. K. Bulik-Sullivan et al., 2015). All variants in the MHC region (GRCh37: 6:28,477,797–33,448,354) were removed prior to the analysis. Heritability estimates were calculated for subregions from the FreeSurfer segmentation and the Hofer and Frahm segmentation. Genetic correlations were calculated for total CC volume, each subregion and relevant traits which had indicated CC alterations in prior imaging studies and which had publicly available GWAS. Genetic correlations were calculated for: attention deficit hyperactivity disorder (Stergiakouli *et al*., 2012), autism spectrum disorder (PGC ASD, 2017), BD (Mullins *et al*., 2021), major depressive disorder (MDD) (Wray *et al*., 2018), schizophrenia (SCZ) (Liu *et al*., 2021), the PGC cross-disorder GWAS (Pouget *et al*., 2019), cigarettes per day, drinks per week, smoking initiation, smoking cessation (Liu *et al*., 2019) and cannabis use (Pasman *et al*., 2018).

## Results

### Genome-wide association analyses

We found 70 unique genetic loci with distributed associations across the CC, using MOSTest. The polygenic architecture of the CC is shown in Figure 1A, illustrating the results of the multivariate analysis. The univariate statistics indicate subregion-specific genetic signals for each of the CC subregions (Figure 1). The loci that showed strongest significant effects from the multivariate analysis showed distributed effects with varying patterns, often exhibiting a gradient from posterior to anterior across CC subregions (Figure 1B). A list of significant SNPs from the univariate GWAS of total CC volume and the volume of each CC subregion can be found in the supplementary material (Supplementary Material, Table 1). Figure 2A displays the results from the univariate GWAS for the volume of each subregion. The results for the GWAS, co-varying for total CC volume can be found in supplementary material, Figure 3.

**Figure 3:**
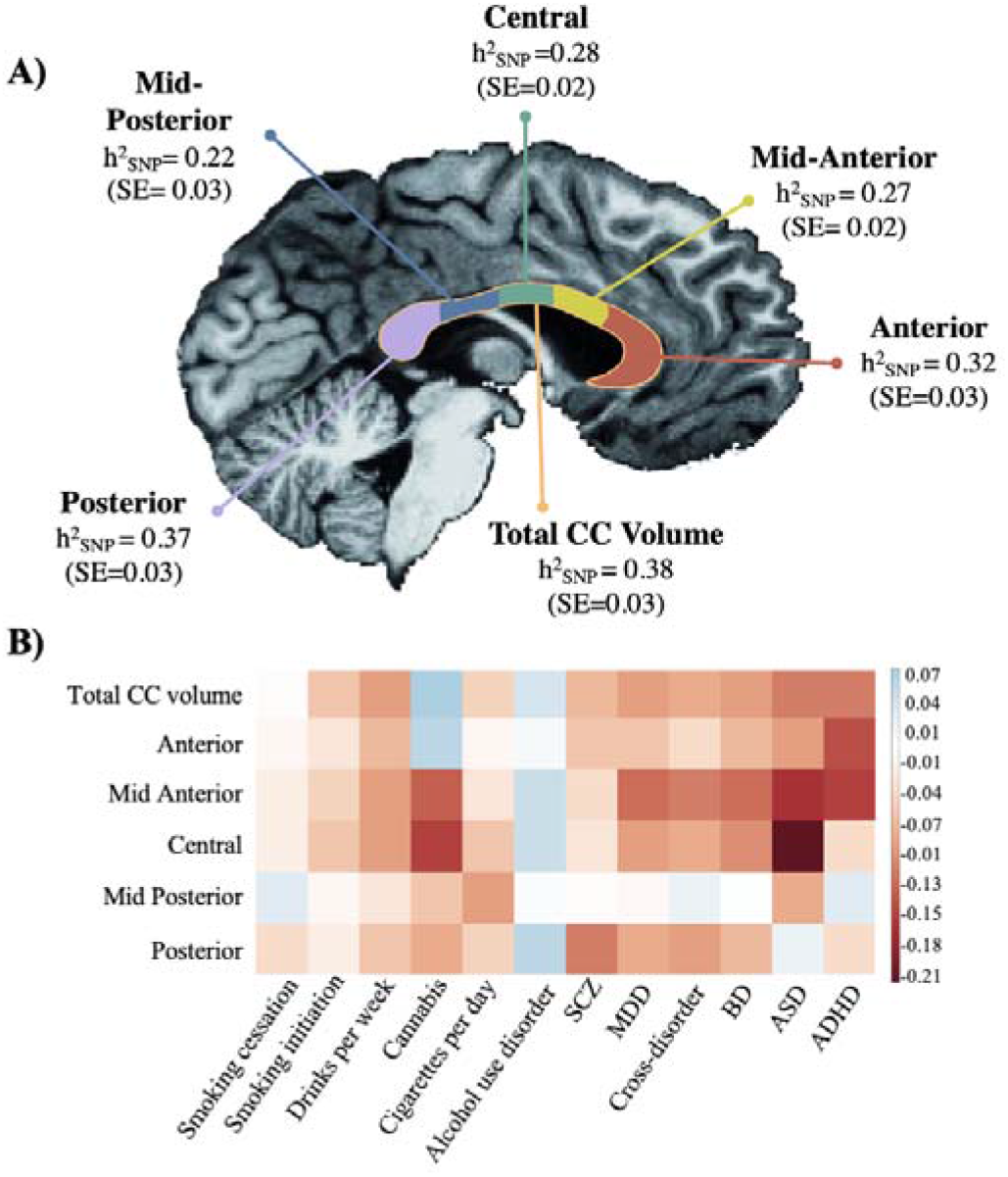
**A) SNP based heritability estimates of total CC volume and volume of the subregions** LDSC was used to estimate the SNP-based heritability of total CC volume and the five subregions. A schematic of the CC and the five subregions, extracted using FreeSurfer, based on the Witelson segmentation. The CC and subregions have been overlaid onto an anatomical reference image. **B) Heatmap of genetic correlations** between CC subregions and relevant psychiatric and substance use traits

Replication of the multivariate results was conducted using MOSTEST-PVS in the 5,286 individuals of non-European ancestry, for the FreeSurfer segmentation. Of the 70 significant, lead SNPs in the multivariate analysis of the European sample, only 33 were available in the individuals of non-European ancestry. Of these 33 SNPs, all had the same direction of effect in the group of non-European ancestry. Twenty-two of the 33 SNPs are nominally significant (*p*<0.05) in the replication and six remained significant after correction for multiple testing (Bonferroni, *p*<1.51×10^−3^) (Supplementary Material, Table 2). The results for the multivariate analysis of the Hofer and Frahm segmentation can be found in Supplementary Material, Figure 4.

Independent, lead SNPs from the univariate, European GWAS were extracted and a linear regression was conducted in the non-European individuals. Of the 195 lead SNPs from the univariate GWAS of total CC and the univariate GWAS of each subregion, 140 were present in the non-European replication sample. Fifty-four of the SNPs are nominally significant (p<0.05) and 5 SNPs remained significant after correction for multiple testing (Bonferroni, *p*<0.05/140=3.57×10^−4^). Consistent directions of allelic effect were observed in the non-European analysis when compared to the European lead SNPs, with 89% of the SNPs having the same direction of effect (Supplementary Material, Table 2). (Supplementary Material, Table 2).

### Functional annotation

Significant gene sets for the GWAS of each subregion and total CC are listed in Supplementary Material, Table 3. Gene sets identified for the anterior, mid-anterior and central subregion overlapped significantly with previous GWAS of bipolar disorder, alcohol use disorder, Alzheimer’s dementia, loneliness and neuroticism (Nagel *et al*., 2018; Turley *et al*., 2018; Jansen *et al*., 2019; Kranzler *et al*., 2019; Stahl *et al*., 2019). Gene sets identified for the posterior subregion overlapped with multiple hippocampal phenotypes and implicated cytoskeleton organization (van der Meer, Rokicki, *et al*., 2020). Manhattan plots from the gene-based association tests, conducted through FUMA, can be found in Supplementary Material, Figure 4. PheWAS, conducted with Open Target, implicated kidney and liver function (Supplementary Material, Table 4).

### SNP-based heritability and Genetic Correlation

SNP-based heritability was estimated using LDSC for both the FreeSurfer and the Hofer and Frahm segmentation (B. Bulik-Sullivan et al., 2015; B. K. Bulik-Sullivan et al., 2015). The heritability of total CC volume was estimated to be 0.38 (SE=0.03) in the FreeSurfer segmentation, with the posterior subregion having the highest heritability estimate (h^2^_SNP_=0.37, SE=0.03) and the mid-posterior subregion having the lowest (h^2^_SNP_=0.22, SE=0.03). The heritability estimates for the Hofer and Frahm segmentation showed the mid-anterior subregion having the highest heritability (h^2^_SNP_=0.32, SE=0.03) and the mid-posterior subregion having the lowest (h^2^_SNP_=0.26, SE=0.02). All heritability estimates for the FreeSurfer segmentation are illustrated in Figure 3A and a full table of heritability estimates for the Hofer and Frahm segmentation can be found in the Supplementary Material, Table 5.

Using LDSC, we found significant genetic correlations between total CC volume, bipolar disorder (BD, *r*_*g*_=−0.09, SE=0.03; *p*=5.9×10^−3^) and drinks per week (*r*_*g*_=−0.09, SE=0.02; *p*=4.8×10^−4^). Additionally, we found the significant genetic correlations between volume of the mid anterior subregion BD (*r*_*g*_=−0.12, SE=0.02; *p*=2.5×10^−4^), major depressive disorder (*r*_*g*_=−0.12, SE=0.04; *p*=3.6×10^−3^), drinks per week (*r*_*g*_=− 0.13, SE=0.04; *p*=1.8×10^−3^) and cannabis (*r*_*g*_=−0.09, SE=0.03; *p*=8.4×10^−3^) (Figure 3B). A full table of these results can be found in Supplementary Material, Table 6.

## Discussion

Several key findings emerged from this work. First, we identified 70 unique genetic loci contributing to CC volume, with distributed associations across the structure, indicating a polygenic architecture. Second, subregions of the CC also have different genetic architecture, providing evidence that the CC and its subregions have both overlapping and unique genetic contributions. Third, we note significant genetic overlap of CC and CC subregions with neuropsychiatric disorders, including BD, MDD, ASD, as well as cannabis and alcohol consumption.

We identified 70 novel loci that were associated with CC volume, and these loci had distributed effects across the five subregions of the CC. This finding suggests that there is a similar biology underlying the development of these different subregions, and that they may share common genetic pathways. By using multivariate analysis, we were able to boost genetic discovery and gain a more comprehensive understanding of the genetic architecture of the CC and its subregions.

Our univariate GWAS indicate that the five subregions making up the CC have both a shared and distinct genetic etiology, with the loci associated with whole CC volume only partially overlapping with those of the individual CC subregions. We identified a number of novel loci associated with CC subregional volume (33 significant loci in the anterior subregion, 23 in the mid anterior, 29 in the central, 7 in the mid posterior and 56 in the posterior subregion). Although there were no SNPs that overlapped with all subregions, rs755857 was significant in each GWAS except for that of the mid-posterior. Although rs755857 has not been previously associated with any other trait, it is associated with gene *PINX1*. Alterations in *PINX1* have been associated with gamma glutamyl transferase and serum alkaline phosphatase levels both of which are used as clinical markers of liver function (Barton *et al*., 2021).

The SNP-based heritabilities of the CC subregions ranged from 0.22 (mid posterior) to 0.37 (posterior), with a SNP-based heritability of 0.28 for total CC volume. These findings indicate that the CC and its subregions have a higher SNP-based heritability compared to subcortical brain volumes such as the amygdala and hippocampus (Satizabal *et al*., 2019; van der Meer, Frei, *et al*., 2020; Mufford *et al*., 2021; Bahrami *et al*., 2022; Ou *et al*., 2023). Our heritability estimates are consistent with previous family reports indicating that the size of the CC and its subregions are under strong genetic influence (Woldehawariat *et al*., 2014). The anterior and posterior subregions have the highest heritability estimates (*h*^*2*^_*snp*_ =0.32 and *h*^*2*^_*snp*_ =0.37, respectively), consistent with previous work Woldehawariat (2014), who found the anterior and posterior subregions of the CC to have the highest heritability estimates. Our findings corroborate this pattern of high heritability estimates in the anterior and posterior subregions, and lower estimates in the mid-anterior and mid-posterior subregions.

Gene-set analyses conducted through FUMA indicated a degree of overlap between the subregions, with the exception of the mid-posterior subregion. The mid posterior subregion has a distinct set of significant SNPs, with minimal genetic overlap with the other subregions and no gene sets in common with the other subregions. Additionally, the mid posterior subregion had the lowest number of significant loci and the smallest heritability estimate. This is in line with previous work which shows that the mid posterior subregion is difficult to examine; perhaps due to its smaller size upon extraction, but to date there are no studies that have assessed its heritability in humans. Studies looking at its heritability in other primates, suggest that it has a lower heritability when compared to larger subregions, such as the anterior subregion (Hopkins *et al*., 2022). Although we found no evidence to suggest a relationship between increasing subregion volume and the number of significant loci detected, this remains a possibility. The subregion has projections into the posterior parietal and superior temporal cortex which also have lower heritability estimates (*h*^*2*^_*SNP*_= 0.07, *SE=*0.02; *h*^*2*^_*SNP*_=0.12, *SE*=0.02, respectively) when compared to other cortical regions, which may explain the lower heritability seen for this region (Grasby *et al*., 2020).

We found significant genetic overlap with previous GWAS of BD, alcohol use disorder and ASD. This is consistent with evidence of CC involvement in these conditions (Prunas *et al*., 2018; de Souza *et al*., 2019; Valenti *et al*., 2020). Our genetic correlation results also echoed findings from previous imaging studies. We found significant negative genetic correlation of BD with total CC volume, as well as volumes of the mid anterior and central subregions. Consistently, BD patients have been shown to have significantly smaller CC subregions in comparison to healthy controls (Arnone, Abou-Saleh and Barrick, 2006; Gifuni *et al*., 2017; Prunas *et al*., 2018). Volumetric MRI studies have consistently demonstrated reduced CC volume in individuals with BD and their healthy co-twins, implying that these abnormalities may be related to genetic factors (Thompson *et al*., 2003; Bearden *et al*., 2011). Further, volumetric reductions in the CC have been observed in relatives of patients with BD, showing intermediate volumetric reductions when compared to both controls and BD probands (Walterfang *et al*., 2009; Francis *et al*., 2016). Our findings add additional evidence to previous work, indicating that altered CC volumes in BD may be due to a shared genetic aetiology. Reductions in total CC volume have also been reported in ASD (Frazier and Hardan, 2009; Valenti *et al*., 2020). Additionally, there appears to be a gradient of volumetric reduction whereby the anterior subregions show more pronounced effect and the posterior comparatively less affected (Frazier and Hardan, 2009). Our genetic correlation results are in line with these findings: there was a significant correlation between ASD and the central subregion.

Previous work has also shown that alcohol consumption affects CC morphometry, irrespective of an alcohol use disorder diagnosis (de Souza *et al*., 2019). Here we find significant genetic correlations between the number of drinks per week and total CC, mid anterior and central subregion volumes. Since alterations in the CC are consistently noted in individuals who consume alcohol, our findings suggest that there may be a genetic underpinning to these changes. The exact nature of this relationship warrants further investigation. The top SNP from our multivariate analysis, rs11245366 is associated with the gene *EEF1AKMT2*. PheWAS has found this SNP to be associated with cystatin C, creatine and urate levels (Barton *et al*., 2021). These phenotypes are related to kidney function and are markers of kidney dysfunction. Abnormalities in renal function have been consistently noted in individuals with agenesis of the CC; additionally, alterations in white matter structure are present in individuals with chronic kidney disease (Franco *et al*., 1993; Liu *et al*., 2020). Here we provide preliminary evidence for a shared genetic etiology between these phenotypes. The genetic relationship between the CC and renal phenotypes provides an interesting new path for future investigation (Fan *et al*., 2019; González-Reimers *et al*., 2019).

Some limitations deserve emphasis. First, our study was limited to individuals residing in the United Kingdom and consisted largely of individuals of European ancestry. We did however, replicate our findings in an independent sample of non-European ancestry. Future genetic work may benefit additional analyses conducted in more diverse cohorts. Second, the age range of individuals in the UK biobank is limited to those above 45 years old. Therefore, it would be important to replicate these findings in a younger sample to ensure generalizability. Third, the comparison of our results with the literature is challenging due to differences in approach to subregion divisions. Although we implemented two segmentation protocols with consistent results, a robust CC segmentation is needed to improve replicability of research in this field.

In conclusion, our results demonstrate that the CC has a polygenic architecture with multiple genes contributing to CC volume, but also suggest that distinct genetic factors are involved in the development of anterior and posterior subregions, consistent with their divergent functional specialization. The pheWAS findings emphasize the importance of liver and kidney function in CC volume. We provide the first SNP-based evidence that CC and subregion volumes are heritable. Genetic correlations of CC and subregions volumes with specific psychiatric traits is noteworthy, and deserving of further investigation.

### Note in Support

The work in this paper was at the World Congress of Psychiatric Genetics (Florence, 2022), the American Society of Human Genetics (Los Angeles, 2022) and at the International Congress of Human Genetics (Cape Town, 2023) (Campbell *et al*., 2022). While in the process of uploading this paper for submission, we noted a paper by Chen et al., 2023 (Chen *et al*., 2023). This publication takes a similar approach, albeit with some differences in methods and results, so providing additional support for our findings.

